# Sensitivity of unconstrained quantitative magnetization transfer MRI to Amyloid burden in preclinical Alzheimer’s disease

**DOI:** 10.1101/2024.04.15.24305860

**Authors:** Andrew Mao, Sebastian Flassbeck, Elisa Marchetto, Arjun V. Masurkar, Henry Rusinek, Jakob Assländer

## Abstract

Magnetization transfer MRI is sensitive to semi-solid macromolecules, including amyloid beta, and has previously been used to discriminate Alzheimer’s disease (AD) patients from controls. Here, we fit an unconstrained 2-pool quantitative MT (qMT) model, i.e., without constraints on the longitudinal relaxation rate 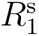 of semi-solids, and investigate the sensitivity of the estimated parameters to amyloid accumulation in preclinical subjects. We scanned 15 cognitively normal volunteers, of which 9 were amyloid positive by [^18^F]Florbetaben PET. A 12 min hybrid-state qMT scan with an effective resolution of 1.24 mm isotropic and whole-brain coverage was acquired to estimate the unconstrained 2-pool qMT parameters. Group comparisons and correlations with Florbetaben PET standardized uptake value ratios were analyzed at the lobar level. We find that the exchange rate and semi-solid pool’s 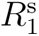 were sensitive to the amyloid concentration, while morphometric measures of cortical thickness derived from structural MRI were not. Changes in the exchange rate are consistent with previous reports in clinical AD, while changes in 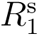 have not been reported previously as its value is typically constrained in the literature. Our results demonstrate that qMT MRI may be a promising surrogate marker of amyloid beta without the need for contrast agents or radiotracers.

## 1 Introduction

Conformational abnormalities in the amyloid β (Aβ) protein are one of the defining pathological hallmarks of Alzheimer’s disease (AD) (Jack et al., 2018; Thal et al., 2002). In the extracellular space, 40-42 amino acid Aβ peptide fragments cleaved from amyloid precursor protein, known as Aβ_40_ and Aβ_42_, form soluble aggregates known as oligomers, which further aggregate into insoluble fibrils and ultimately plaques (Karran et al., 2011). It is believed that several species of Aβ aggregates have neurotoxic or inflammatory effects (Heppner et al., 2015) that ultimately lead to tau hyperphosphorylation (Oddo et al., 2006), neurodegeneration, and cognitive symptoms (Bennett et al., 2004), though the exact mechanisms are unclear. Aβ accumulation begins in the neocortex (Fantoni et al., 2020), even in clinically asymptomatic subjects, and can occur decades prior to the possible symptomatic onset of dementia (Jack et al., 2010). Identifying individuals with a substantial amyloid load is necessary for many clinical reasons, including the specific diagnosis of AD versus other dementias (Jack et al., 2018), and to study the impact of new disease-modifying therapies—such as emerging anti-amyloid immunotherapeutics— which are being used to treat early stages of AD and increasingly being studied in preclinical patient populations (Yadollahikhales & Rojas, 2023).

The gold standard for noninvasive *in vivo* amyloid assessment is Positron Emission Tomography (PET) (Chapleau et al., 2022; Johnson et al., 2013; Rabinovici et al., 2023). The second generation of amyloid PET tracers includes [^18^F]Florbetaben, which has been demonstrated to have high sensitivity for fibrillar amyloid (Syed & Deeks, 2015). While cerebrospinal fluid (CSF) Aβ biomarkers are similarly sensitive and lumbar punctures confer additional access to tau biomarkers (Husźar et al., 2024; Palmqvist et al., 2015), intravenous radiotracer injection followed by PET imaging is less invasive. Additionally, PET offers spatial localization of the Aβ signal which may confer the ability to detect regional amyloid depositions before pathological changes occur in the global neocortical signal. However, PET has several drawbacks, including cost, the need for specialized equipment, availability, limited spatial resolution, off-target binding, and ionizing radiation exposure.

An alternative, magnetic resonance imaging (MRI)-based technique that may be directly sensitive to the accumulation of extracellular protein deposits, such as Aβ aggregates, is known as quantitative magnetization transfer (qMT) (Henkelman et al., 1993). MT methods sensitize the MRI signal to a “semi-solid” spin pool consisting of protons bound in large macromolecules such as lipids (e.g., myelin) and proteins (e.g., both Aβ_40_ and Aβ_42_ aggregates), which exchange with protons bound in the usual “free water” pool. MT’s sensitivity to the accumulation of insoluble Aβ plaques was previously demonstrated in transgenic mice (Bigot et al., 2014; Pérez-Torres et al., 2014; Praet et al., 2016), and *in vivo* studies using *quantitative* MT—which disentangles the biophysical contributions to the MT contrast—suggest that the “forward magnetization exchange rate” is the most discriminatory qMT biomarker for classifying AD versus controls and the conversion from amnestic mild cognitive impairment (MCI) to AD (Duan et al., 2022; Giulietti et al., 2012; Makovac et al., 2018).

It is unclear from prior qMT studies whether the observed differences in the “forward exchange rate” arise from changes in the exchange rate (e.g., due to the insolubility of Aβ plaques) or the macromolecular pool size (e.g., from neurodegeneration). Additionally, prior studies usually constrain the value of the difficult-to-estimate semi-solid pool’s longitudinal relaxation rate 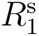. In this work, we quantify all parameters of the unconstrained 2-pool qMT model using a tailored *hybrid-state* pulse sequence (Assländer, Mao, et al., 2024). Furthermore, due to previous works’ focus on clinically-diagnosed MCI or AD (Duan et al., 2022; Giulietti et al., 2012; Makovac et al., 2018), it is unclear whether *preclinical* AD pathology can be detected with qMT biomarkers. This distinction is important because Aβ has been shown to accumulate well before the clinical manifestations of dementia (Jack et al., 2010). Therefore, we focus on the preclinical population in this study. Our central hypothesis is that amyloid accumulation in the preclinical population is detectable by qMT biomarkers—including 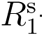 —due to the distinct biochemical properties of Aβ plaques.

## 2 Methods

### 2.1 Magnetization Transfer Model

We use the unconstrained qMT model (Figure 1) presented in Assländer, Mao, et al. (2024), which describes the Bloch-McConnell equations (McConnell, 1958) of the 2-pool spin system (Henkelman et al., 1993):

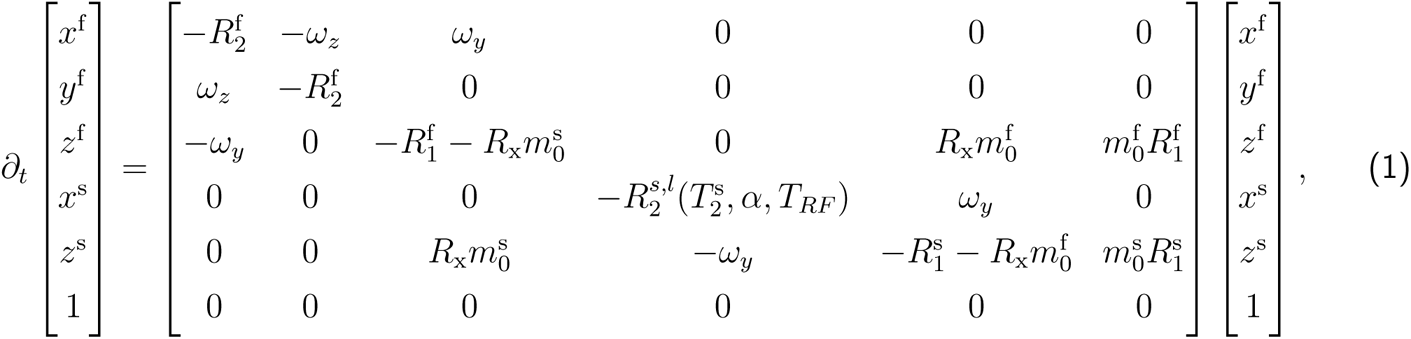

where *ω_z_* is the off-resonance frequency and *ω_y_* is the Rabi frequency of the radiofrequency (RF) pulse which has a flip angle *α* and duration *T_RF_* . The magnetization of protons bound in free water (super-script ^f^ ) is described in Cartesian coordinates by *x*^f^ *, y*^f^ *, z*^f^ . The free pool’s magnetization exchanges at the rate *R*_x_ with the semi-solid spin pool (superscript ^s^), whose magnetization is captured in Cartesian coordinates by *x*^s^*, z*^s^. Each pool has its own fractional size 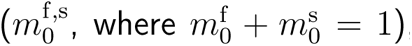, longitudinal 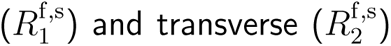 relaxation rates (inverse of times *T*_1_,_2_), amounting to six core parameters: 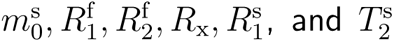. The non-exponential decay characteristics of the semi-solid pool in the brain’s parenchymal tissue are characterized by the super-Lorentzian lineshape (Morrison et al., 1995), where we use the time 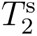 (instead of the rate) for consistency with the qMT literature. These decay characteristics are incorporated in the Bloch-McConnell equation with the generalized Bloch model (Assländer et al., 2021). For computational efficiency, we approximate the non-exponential decay characteristics with an effective exponential decay using the *linearized* relaxation rate 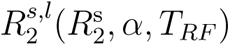 that results in the same magnetization at the end of an RF pulse with the flip angle *α* and pulse duration *T_RF_* . Without loss of generality, we neglect *y*^s^ assuming *ω_x_* = 0 and 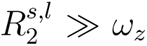. More details regarding the generalized Bloch model are described in Assländer et al. (2021). Note the slight differences with respect to quantities often referenced in the MT literature, including the pool-size ratio 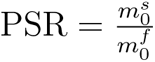 (Henkelman et al., 1993) where 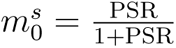, and the “forward exchange rate,” which is the product 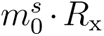. For the latter, many previous studies could not estimate the parameters 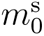 and *R*_x_ separately due to limitations of the employed encoding strategies and signal models (Ramani et al., 2002), which is overcome here by the use of a tailored hybrid-state pulse sequence.

**Figure 1:**
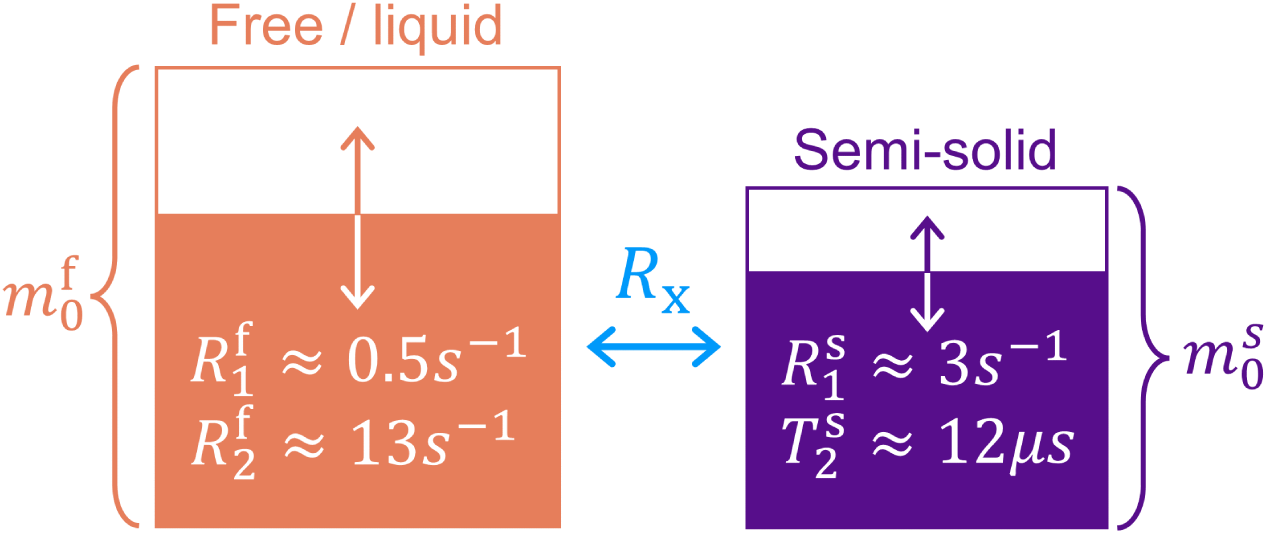
2-pool quantitative magnetization transfer model (Assländer, Mao, et al., 2024; Henkelman et al., 1993). The red-colored pool models “free water” protons with fraction 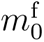, while macromolecule-bound “semi-solid” protons with fraction 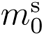 are colored purple 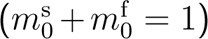. Each pool’s longitudinal and transverse relaxation rates (reciprocal of the times) 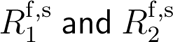, respectively, are modeled separately (most previous studies constrain the value of 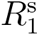). After saturation of one pool’s longitudinal magnetization, relaxation and exchange with the other pool modulate each pool’s magnetization as visualized by the arrows and partially colored boxes. The exchange rate is denoted by *R*_x_.

The highly restricted motion of macromolecules leads to an ultrashort 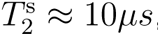, which prevents direct detection of this pool with the typical echo times achievable on clinical MRI scanners. Hence, the semisolid pool can be detected on clinical MRI scanners only indirectly via its exchange with the free pool, impeding the estimation of 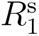. Consequently, authors have typically assumed 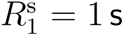 (Giulietti et al., 2012; Henkelman et al., 1993; Makovac et al., 2018). In this work, we overcome these limitations by utilizing a hybrid-state sequence (Assländer, Mao, et al., 2024; Assländer et al., 2019) (more details in Section 2.3). We recently demonstrated *in vivo* the ability to voxel-wise quantify 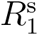 (Assländer, Mao, et al., 2024), which takes on significantly smaller values than 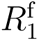 (Helms & Hagberg, 2009; Manning et al., 2021; Samsonov & Field, 2021; van Gelderen et al., 2016). We hypothesized that eliminating this constraint on 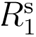 increases qMT’s sensitivity to changes in the semi-solid pool’s biophysical properties arising from Aβ accumulation.

### 2.2 Study Participants

We recruited 15 cognitively normal subjects (Clinical Dementia Rating^®^ of 0) from New York University’s AD Research Center (ADRC) cohort of community-dwelling elderly adults. All subjects had received amyloid PET scans within the preceding 34 months (mean *±* standard deviation 17*±*9 months). Six individuals were considered Aβ*−* by our ADRC’s standardized uptake value ratio (SUVR) threshold, with the following demographics: three female, three white, age 72.6*±*4.5 years (mean *±* standard deviation), Montreal Cognitive Assessment (MoCa) scores 27.3*±*2.2, one *APOE* -ε4 and two *APOE* -ε2 carriers. The remaining nine subjects were Aβ+, with the demographics: five female, seven white, age 75.6*±*6.5 years, MoCa 27.4*±* 2.4, six *APOE* -ε4 carriers. All subjects provided written informed consent for the studies described below, in agreement with the requirements of the New York University School of Medicine Institutional Review Board.

### 2.3 Imaging Protocol

All subjects received 300 MBq (8.1 mCi) of [^18^F]florbetaben (FBB) tracer (Life Molecular Imaging, Totowa, NJ) intravenously over 15 seconds, followed by a 12-cc saline flush. Syringes were assayed pre- and post-injection. Subjects rested in the injection room to achieve brain equilibration of the tracer before being positioned in the scanner. Amyloid PET scans were acquired on a 3 Tesla Biograph mMR PET/MRI system (Siemens Healthineers, Germany) 90–120 minutes post-injection. Structural MRI was also obtained for registration to images from the next session.

In a second session 17*±*9 months (no more than 34 months) later, each subject underwent a 24 minute MRI exam on a 3 Tesla Prisma MRI scanner (Siemens Healthineers, Germany) using a 32-channel head coil. Our experimental whole-brain qMT technique used a hybrid-state sequence (Assländer et al., 2019) optimized for quantifying MT parameters with a nominal 1 mm isotropic resolution (effectively 1.24 mm isotropic accounting for 3D radial koosh-ball sampling of only the in-sphere of a 1 mm k-space cube) across a 256*×*256*×*256 mm FOV in 12 minutes (Assländer, Mao, et al., 2024). The hybrid-state sequence is similar to an inversion-recovery balanced steady-state free precession (bSSFP) sequence (Bieri & Scheffler, 2013; Carr, 1958) in that it uses fully balanced gradient moments per repetition time, but it also incorporates a smoothly varying flip angle and RF pulse duration between repetition times (Assländer et al., 2019) that was optimized for the encoding of the qMT parameters (Assländer, Mao, et al., 2024). However, we note that the RF pattern was optimized for measuring demyelination in white matter rather than tailored specifically for the detection of Aβ in the cortex. k-Space readout was performed with radial koosh ball trajectory, where the direction of the 3D radial spokes was distributed across the unit sphere using a 2D golden means pattern (Chan et al., 2009), reshuffled to minimize eddy current artifacts (Flassbeck et al., 2024). More details about the hybrid-state sequence—including the encoding mechanisms and its numerical optimization for quantifying the unconstrained qMT parameters—can be found in Assländer, Mao, et al. (2024).

3D Magnetization-Prepared Rapid Acquisition Gradient-Recalled Echo (MPRAGE) (Brant-Zawadzki et al., 1992; Mugler & Brookeman, 1990) and T2-weighted Fluid-Attenuated Inversion-Recovery (FLAIR) (Hajnal et al., 1992) images were also acquired with 1 mm isotropic resolution. Both sequences were GRAPPA 2x accelerated, where the MPRAGE used a TE/TR/TI (echo time, repetition time, and inversion time) of 2.98 ms/2.3 s/900 ms for 5 min 30 s of scan time and the FLAIR used a TE/TR/TI of 392 ms/5 s/1.8 s for 5 min 57 s of scan time.

### 2.4 Image Reconstruction

PET reconstruction used the Standardized Centralized Alzheimer’s & Related Dementias Neuroimaging (SCAN) parameters (https://scan.naccdata.org), except modified to use only a single frame of the first 10 mins of data to reduce motion-related artifacts (Koesters et al., 2016): OSEM-3D (Erdogan & Fessler, 1999) with 4 iterations and 21 subsets; 344*×*344*×*127 grid; 2.0 zoom (1.04313*×*1.04313*×*2.03125 mm voxels); all corrections on; post-reconstruction smoothing with a 2 mm full width at half maximum Gaussian kernel. Attenuation correction was performed using MR-based hybrid segmentation and atlas-based algorithm that combines tissue segmentation from a Dixon µ-map with a superimposed, co-registered, skull atlas-derived bone compartment (Koesters et al., 2016).

For the qMT sequence, we performed self-navigated motion correction retrospectively with a temporal resolution of 4 s based on low-resolution reconstructions (4 mm isotropic) in a subspace optimized for contrast between the parenchyma and CSF (Flassbeck et al., 2024; Kurzawski et al., 2020). From the corrected k-space data, we reconstructed 15 coefficient images corresponding to a low-rank representation of the MRI signal’s temporal dynamics using a subspace modeling approach (Assländer et al., 2018; Liang, 2007; McGivney et al., 2014; Tamir et al., 2017). The subspace was spanned by singular vectors computed from a dictionary of signals (or fingerprints) and, additionally, their orthogonalized gradients, which maximizes the information needed to stably perform parameter estimation for complex pulse sequences (Mao et al., 2023). The reconstruction also utilized sensitivity encoding (Pruessmann et al., 2001; Sodickson & Manning, 1997), with coil sensitivities estimated using ESPIRiT (Uecker et al., 2014), and locally low-rank regularization to minimize undersampling artifacts and noise (Lustig et al., 2007; Trzasko & Manduca, 2011; T. Zhang et al., 2015). The reconstruction was implemented using the optimal iterative soft thresholding algorithm (Jang et al., 2023) in *Julia* based on publicly available source code (see Section 5). More details about the reconstruction can be found in Tamir et al. (2017) and Assländer et al. (2018).

### 2.5 qMT Model Fitting

Following image reconstruction, we estimated maps of the six qMT parameters 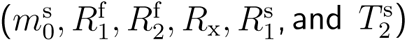 by voxel-wise fitting the unconstrained qMT model to the reconstructed coefficient images. For computational efficiency, we used a neural network-based estimator. This network, tailored for our sequence, takes the 15 complex-valued coefficients, split into real and imaginary parts, as inputs. The architecture was similar to the design described in Figure 2 of X. Zhang et al. (2022): 11 fully connected layers with batch normalization and skip connections with a maximum layer width of 1024. The network incorporated a data-driven correction for *B*_0_ and 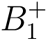 inhomogeneities as described in (Assländer, Gultekin, et al., 2024), and was trained explicitly to minimize the bias that is typically introduced when assuming a distribution for the simulated training data (Mao et al., 2024). Example parameter maps for an Aβ+ subject are shown in Figure 2.

**Figure 2:**
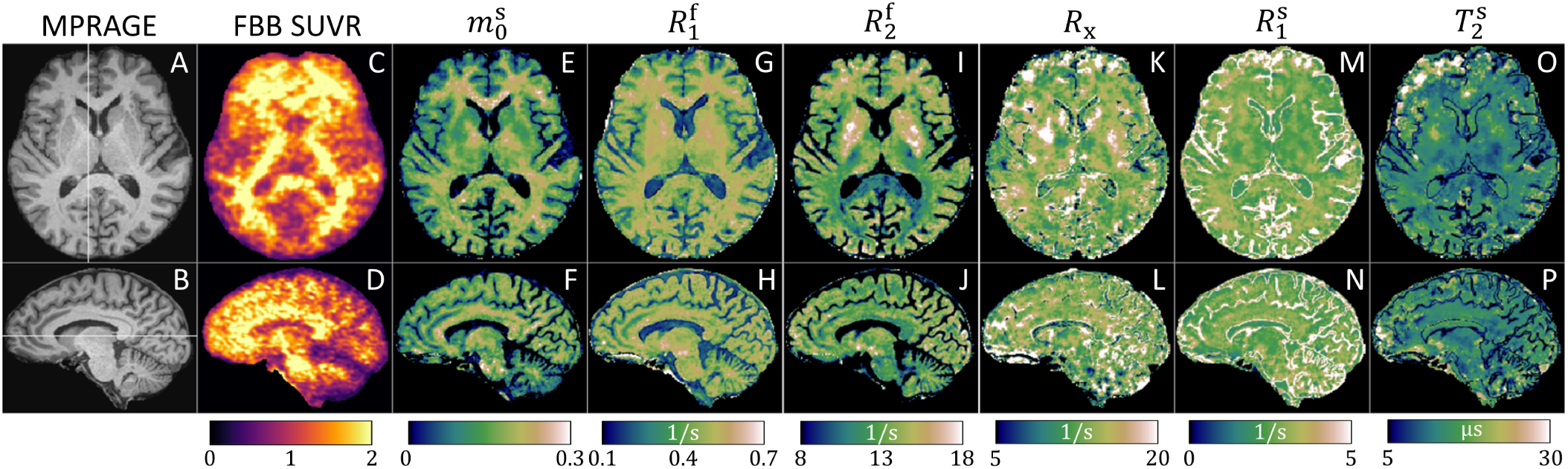
Exemplary MPRAGE (A–B), [^18^F]Florbetaben (FBB) PET SUVR (C–D), and quantitative magnetization transfer maps (E–P) in a cognitively normal Aβ+ subject. The six qMT parameters are the semi-solid pool size 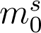, the relaxation rates 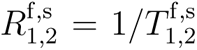 (where the superscripts ^f^ and ^s^ denote the free and semi-solid spin pools, respectively), and the exchange rate *R*_x_. The MPRAGE is used for Freesurfer-based cortical parcellation and calculation of cortical thicknesses. The FBB SUVR images are used to compare the qMT parameters to a relative concentration of Aβ.

### 2.6 Image Processing

The MPRAGE, FLAIR, and FBB images were skull-stripped (Hoopes et al., 2022) and rigid-body registered (Reuter et al., 2010) to the estimated 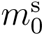 maps (which has the most similar contrast to the MPRAGE amongst the qMT parameters) using the “mri synthstrip” and “robust register” commands, respectively, in Freesurfer v7.4.1 (Fischl, 2012). The qMT parameters were chosen as the common reference frame to avoid interpolating the non-linearly processed qMT maps. Cortical and subcortical segmentation, cortical parcellation, volume, and thickness values were then computed using Freesurfer’s “recon-all” command (Fischl, 2004; Fischl et al., 2002). Volumes were normalized by the estimated total intracranial volume, and the average global cerebellar FBB value was used for normalization to compute SUVRs.

Because the cortices are only 2–3 mm thick compared to the 1.24 mm effective isotropic resolution, and since the quantification of the several qMT parameters is unstable in the CSF (where the macromolecular pool size 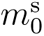 approaches zero), cortical analyses of our qMT parameter maps are highly susceptible to partial volume effects. To mitigate this issue, we adopt a conservative approach of sampling the qMT and FBB SUVR values at the surface corresponding to 50% of the cortical depth using Freesurfer’s “mri vol2surf” function. To avoid resampling the non-linearly processed qMT data and minimize linear interpolation error when using the Freesurfer tools, we applied the following procedure. First, we sinc interpolated (Oppenheim & Willsky, 1996) the reconstructed coefficient images onto a 5*×* finer grid (i.e., 0.2*×*0.2*×*0.2 mm voxels). Then, we used “mri vol2surf” with trilinear interpolation to sample the coefficient images at 50% of the cortical depth before applying the neural network to estimate the qMT parameter maps on the interpolated surface.

### 2.7 Lobar Analysis

Due to small sample sizes, we perform a lobar-level analysis by grouping the Desikan-Killiany cortical ROIs (Desikan et al., 2006) into the four primary cortical lobes (frontal, parietal, temporal, and occipital) using the schema suggested in the Appendix of Klein and Tourville (2012). Each subject’s average measurement (qMT parameter, FBB SUVR, or thickness) per cortical lobe and hemisphere is estimated from the constituent ROIs using an inverse-variance weighting (Kay, 1993) based on each ROI’s sample variance. Prior to all statistical analyses, we manually excluded four ROIs (for all measurements) where we identified substantial artifacts in *R*_x_ across multiple subjects, likely caused by subcutaneous fat (see Section 4.2): the rostral anterior cingulate, precentral, postcentral, and superior parietal gyri. We also excluded three ROIs (for all measurements) superior to the frontal sinus that exhibited bSSFP-like banding artifacts (Bieri & Scheffler, 2013) in the qMT parameters (Figure 2L): the medial and lateral orbitofrontal cortices and temporal pole.

We also consider a synthetic lobe based on the “signature of AD-related cortical thinning” described in Dickerson et al. (2011), which defines a group of cortical regions identified as being the most vulnerable to thinning in a large cohort of cognitively normal individuals who later developed AD dementia. From the Desikan-Killiany atlas, we chose the following ROIs that most closely matched those described in Dickerson et al. (2011) to compute the AD-signature summary measure: the entorhinal cortex, temporal pole, inferior temporal gyrus, angular gyrus, supramarginal gyrus, superior parietal cortex, precuneus, middle frontal gyrus, and superior frontal gyrus.

### 2.8 Statistical Analysis

We used the non-parametric Mann-Whitney *U* test (Fay & Proschan, 2010) to compare measurements between the Aβ*−* and Aβ+ groups. We considered the *p <* 0.05, Bonferroni-corrected *p <* 0.01 (accounting for the five cortical “lobes” considered) and *p <* 0.007 (accounting for six subcortical and global WM ROIs) significance levels, where the latter two were used to account for the family-wise error rate (Dunn, 1961) in the group comparisons shown in Figures 3–4. Effect sizes were quantified using Hedge’s *g* (Hedges, 1981), where we considered 0 *≤ |g| <* 0.5 “small,” 0.5 *≤ |g| <* 0.8 “medium,” 0.8 *≤ |g| <* 1.2 “large,” and *|g| ≥* 1.2 “very large” (Sawilowsky, 2009). A positive effect size (*g >* 0) was defined as measurements that were larger in the Aβ+ group. We used Pearson’s correlation coefficient to evaluate the association of our measurements with amyloid burden, again using the *p <* 0.05 significance level. All statistical analyses were performed using the *HypothesisTests.jl Julia* package.

**Figure 3:**
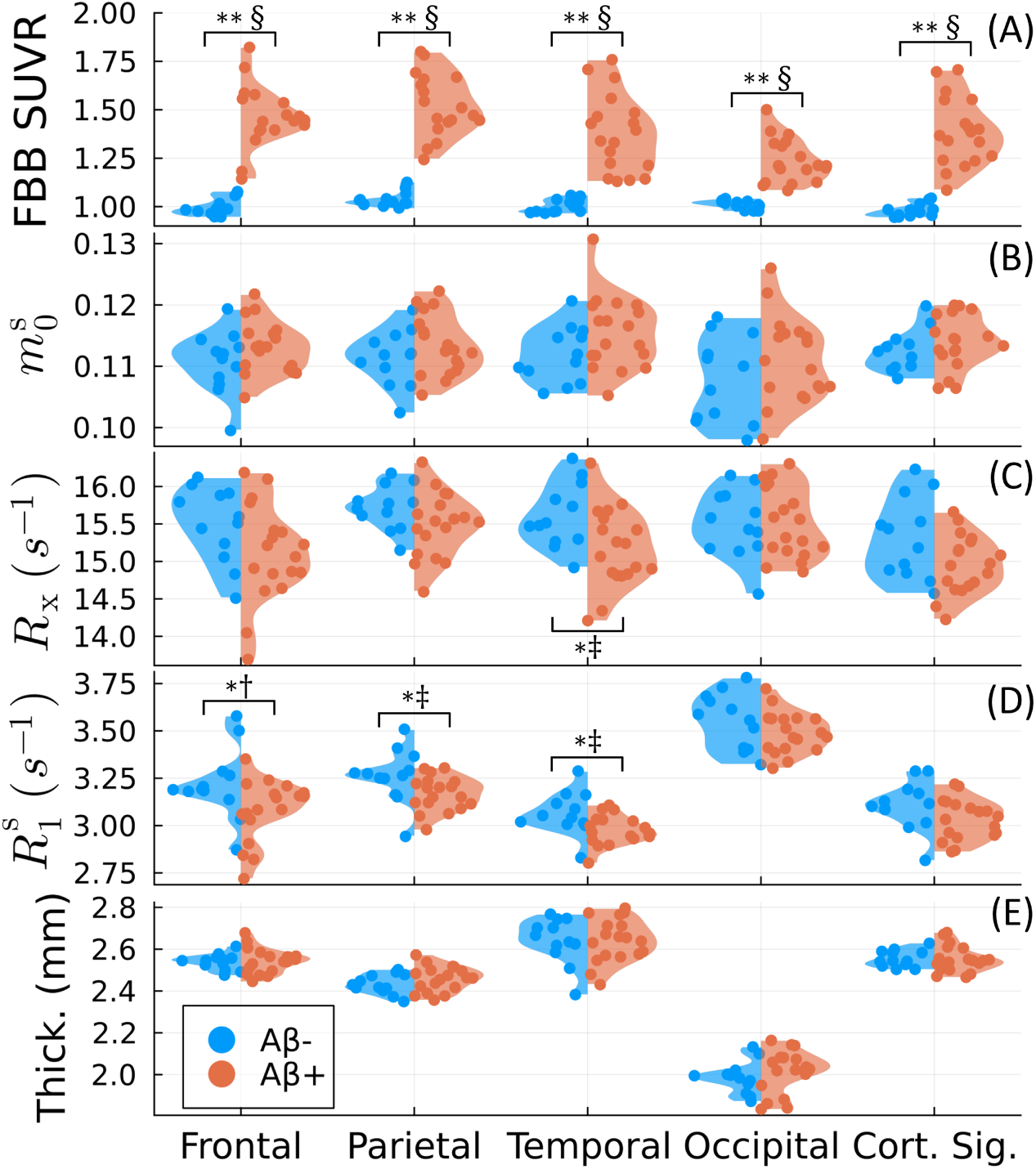
Group comparison for the four cortical lobes and the “cortical signature of thinning” (Dickerson et al., 2011). [^18^F]Florbetaben (FBB) SUVR (A), three qMT parameters (the macromolecular pool size 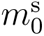 (B), the magnetization exchange rate *R*_x_ (C), and the macromolecular pool’s longitudinal relaxation rate 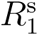 (D)), and cortical thickness (E) are compared between amyloid negative (Aβ*−*) and positive (Aβ+) groups. *∗* and *∗∗* denote statistical significance for *p* < 0.05 and the Bonferroni-corrected *p* < 0.01, respectively, using the Mann-Whitney *U* test (Fay & Proschan, 2010). †, ‡, and § indicate medium, large, and very large effect sizes using Hedge’s *g* (Hedges, 1981; Sawilowsky, 2009), respectively. The *p* and *g* values are summarized in Supporting Table S1 and given for the remaining qMT parameters in Supporting Table S2.

**Figure 4:**
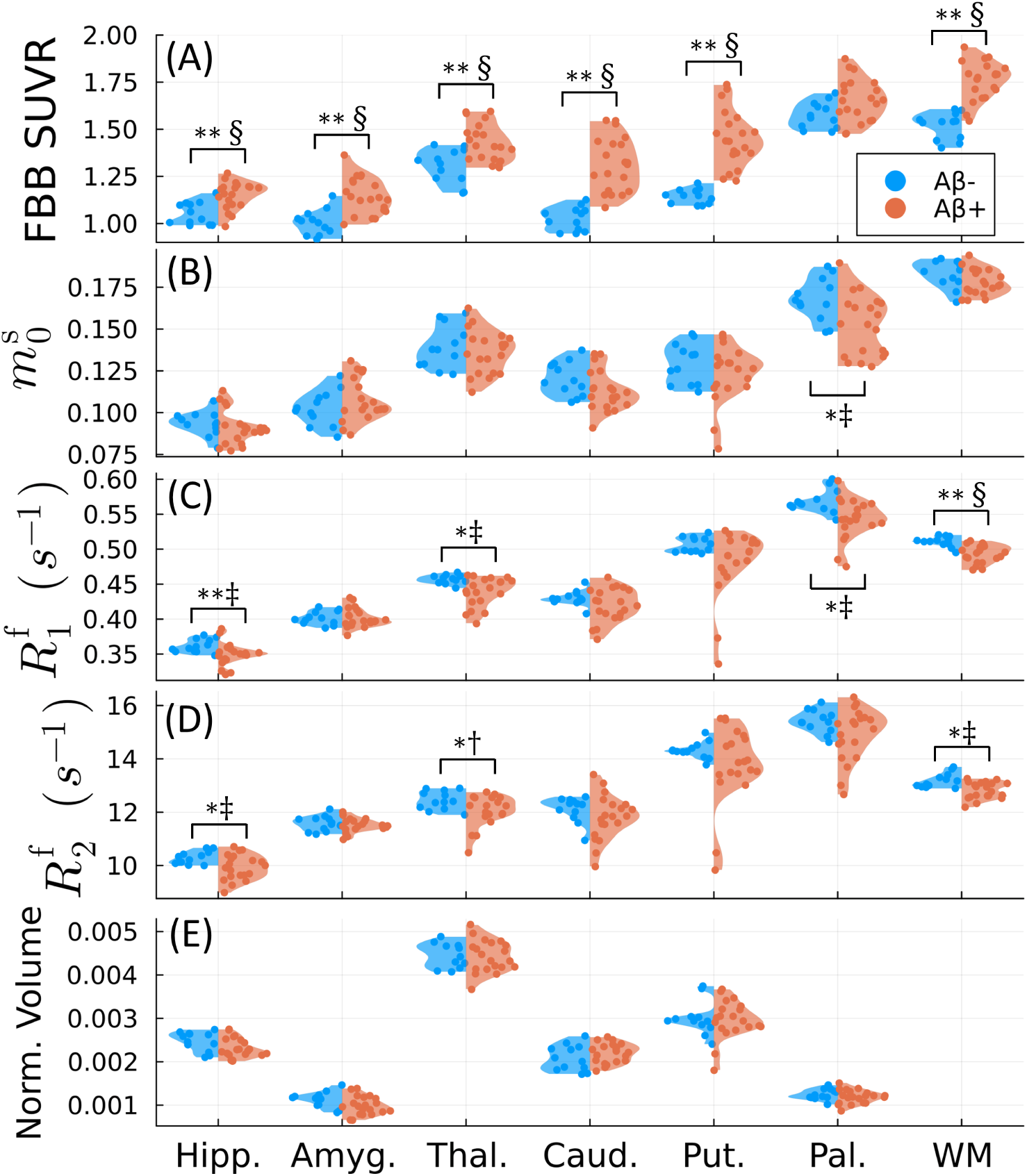
Group comparison for subcortical (hippocampus, amygdala, thalamus, caudate, putamen, pallidum) and global white matter (WM) ROIs. [^18^F]Florbetaben (FBB) SUVR (A), three qMT parameters (the macromolecular pool size 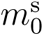, the free pool’s longitudinal relaxation rate 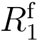 (C), and the free pool’s transverse relaxation rate 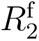 (D)), and volumes (E, normalized by the estimated total intracranial volume) are compared between amyloid negative (Aβ*−*) and positive (Aβ+) groups. Note that the global white matter volumes could not be visualized within the plotted axes, but there were no significant group differences. *∗* and *∗∗* denote statistical significance for *p* < 0.05 and the Bonferroni-corrected *p* < 0.007, respectively, using the Mann-Whitney *U* test (Fay & Proschan, 2010). †, ‡, and § indicate medium, large and very large effect sizes using Hedge’s *g* (Hedges, 1981; Sawilowsky, 2009), respectively. The *p* and *g* values are summarized in Supporting Table S3 and given for the remaining qMT parameters in Supporting Table S4.

## 3 Results

Figure 3 compares the qMT parameters, FBB SUVR, and cortical thickness values between the Aβ*−* and Aβ+ groups. Uniformly across the neocortex, SUVR values were significantly increased in Aβ+ subjects with very large effect sizes, which was expected from our definition of amyloid positivity. Consistently with previous literature reports, the magnetization exchange rate *R*_x_ was significantly decreased in the temporal lobe with a large effect size (*g* = *−*0.86). The macromolecular pool’s longitudinal relaxation rate *R*^s^ was also significantly decreased in the frontal, parietal, and temporal lobes with medium (*g* = *−*0.78), large (*g* = *−*0.81) and large (*g* = *−*0.94) effect sizes, respectively. By contrast, no significant group differences were observed in any lobe for the macromolecular pool size 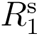, cortical thickness (including for the “AD cortical signature measure”), or the remaining qMT parameters (shown in Supporting Table S2).

We repeated the same analysis for the Freesurfer-defined global white matter and subcortical structures, shown in Figure 4 and Supporting Tables S3–S4. There were no significant differences in the volumes of any subcortical structures (including the hippocampus (Sabuncu et al., 2011)) or total CSF (not shown). However, we observed significant decreases in both free pool relaxation times 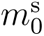 in the global white matter, hippocampus, and thalamus. 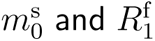 were also significantly decreased in the pallidum.

Figure 5 analyzes the correlation between the qMT parameters or cortical thickness and amyloid concentration (as measured by FBB PET SUVR). Significant Pearson’s correlations occur in similar locations to Figure 3, though with a few differences: there was no significant negative correlation with 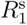 in the parietal lobe, but a significant positive correlation with 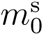 in the temporal lobe. Similarly to Figure 3, no significant correlations were observed between cortical thickness and amyloid concentration in any lobe (neither for the “AD cortical signature measure”).

**Figure 5:**
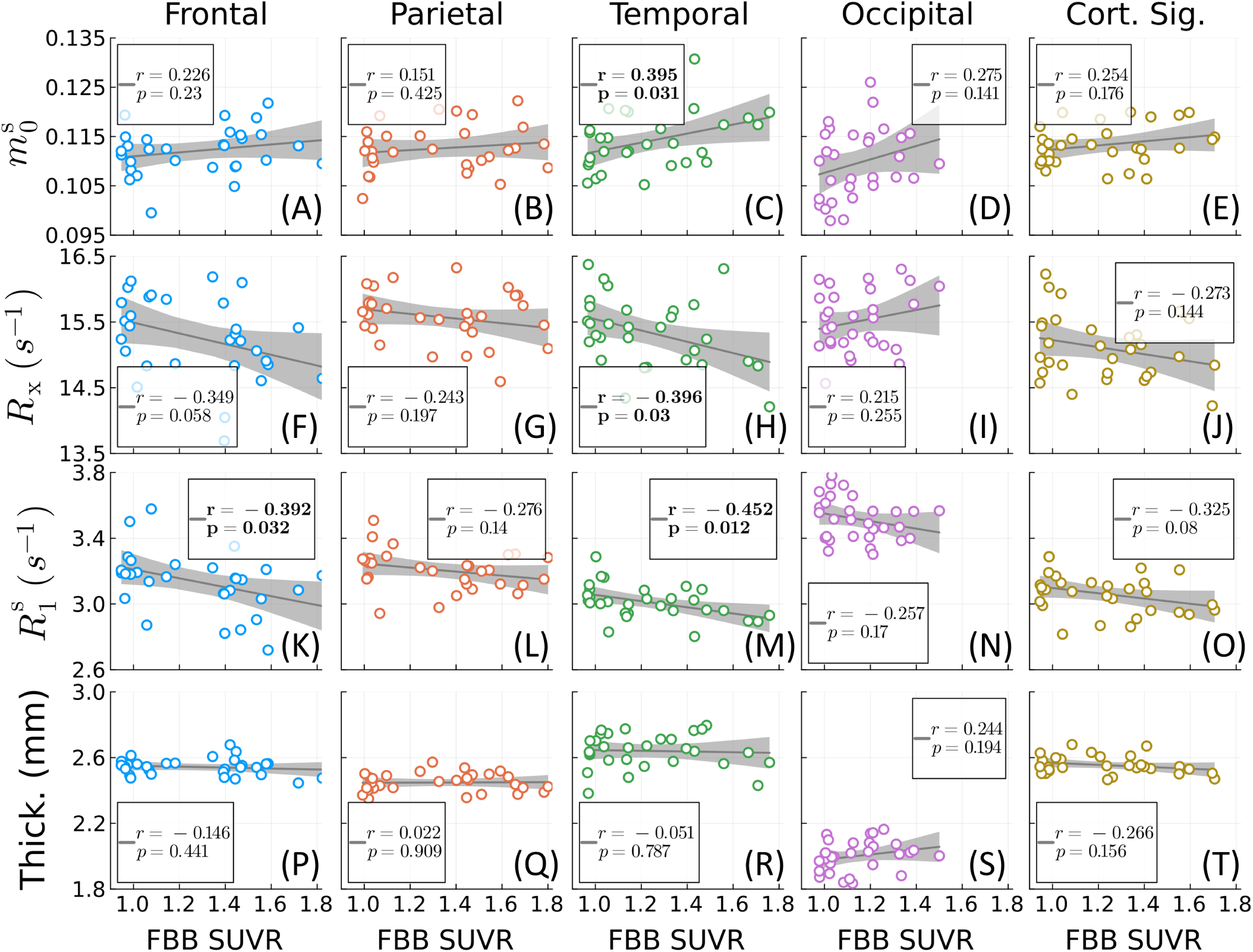
Correlation analysis for the four cortical lobes and the “cortical signature of thinning” (Dickerson et al., 2011). Three qMT parameters—the macromolecular pool size 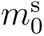 (A–E), the magnetization exchange rate *R*_x_ (F–J), and the macromolecular pool’s longitudinal relaxation rate 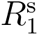 (K–O)—and cortical thickness (P–T) are plotted against amyloid burden as measured by [^18^F]florbetaben (FBB) PET SUVR. Significant Pearson’s correlations (*p* < 0.05) are bolded.

To understand the spatial correspondence between the qMT measurements and FBB SUVR, we visualize the corresponding effect sizes for each cortical ROI overlaid on the Desikan-Killiany atlas in Figure 6. Note that because increases in FBB SUVR as opposed to decreases in 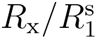 were observed in the Aβ+ group in Figs. 3 and 5, a reversed colorbar is used for FBB SUVR for ease of comparison across the different measurements. As expected, very large positive effect sizes are uniformly observed across the cortex for FBB SUVR. While we do not observe uniformly large *negative* effect sizes for *R*_x_ and 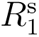 , the ROIs exhibiting (likely spurious) *positive* effects somewhat overlap with the ROIs excluded from the lobar analyses for exhibiting imaging artifacts (e.g., the postcentral and superior parietal gyri in *R*_x_). Importantly, though we do observe small (*g* = *−*0.42) to medium (*g* = *−*0.69) effects suggesting subtle thinning of the right/left entorhinal cortices (Dickerson et al., 2011; Sabuncu et al., 2011), the qMT parameters exhibit higher overall sensitivity to amyloid burden across the entire neocortex.

**Figure 6:**
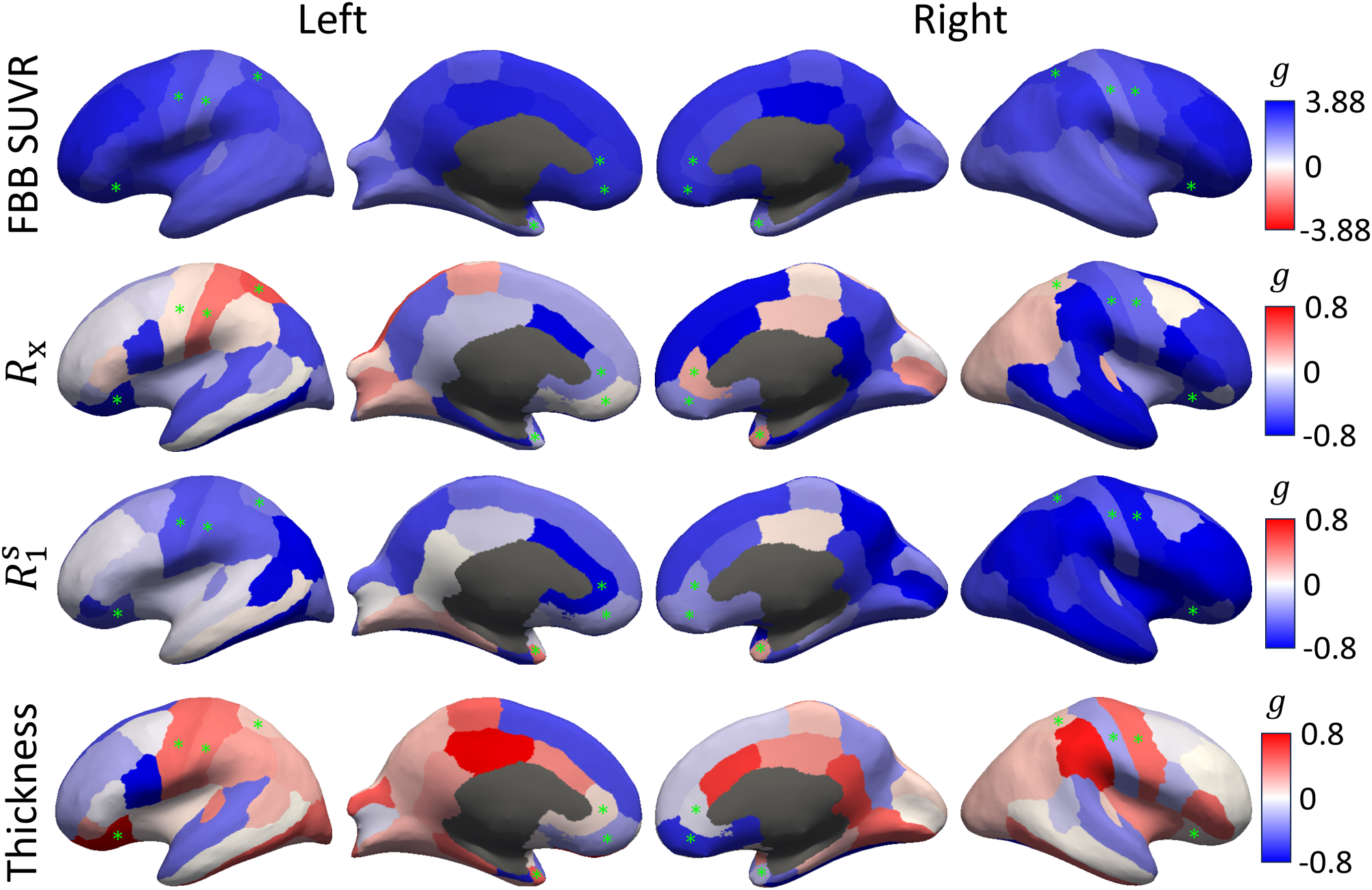
Effect sizes across the neocortex. Hedge’s *g* (Hedges, 1981) comparing Aβ*−* and Aβ+ groups is overlaid on the Desikan-Killiany surface atlas (Desikan et al., 2006) for the magnetization exchange rate *R*_x_, the macromolecular pool’s longitudinal relaxation rate 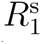, and cortical thickness in comparison to [^18^F]florbetaben (FBB) PET SUVR. A positive effect size (*g >* 0) means measurements are larger in the Aβ+ group, and *|g|* = 0.8 (a “large” effect (Sawilowsky, 2009)) is used as an illustrative cutoff. For FBB SUVR, note the modified cutoffs using the maximal *|g|* value (due to very large positive effect sizes) and the reversed colorbar, used to color the expected direction of effects as blue for all measures (positive for FBB SUVR, negative for *R*_x_, 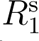, and thickness). The green asterisks denote ROIs excluded from the lobar-level analyses for all measures in Figs. 3–5.

## 4 Discussion

Our study revealed widespread group differences in unconstrained qMT parameters between Aβ+ and Aβ*−* individuals. The largest effect was observed in the semi-solid pool’s longitudinal relaxation rate 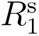. This parameter was previously considered inaccessible and typically constrained. However, using the hybrid-state’s enhanced signal encoding capabilities (Assländer, Gultekin, et al., 2024; Assländer, Mao, et al., 2024; Assländer et al., 2019), we are able to quantify 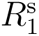 on a voxel-by-voxel basis. Our data suggests that 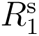 may be a potential biomarker for amyloid pathology preceding morphometric measures of atrophy (Dickerson et al., 2011; Sabuncu et al., 2011). A potential biophysical explanation for this finding could be the slower longitudinal relaxation of spins bound specifically in Aβ plaques relative to other constituents of the macromolecular pool, such as myelin.

We also observed a reduction in *R*_x_ in Aβ+ subjects, which has previously been attributed to the hydrophobicity of Aβ plaques (Chen et al., 2017; Giulietti et al., 2012). This finding aligns with previous reports that the forward exchange rate (i.e., 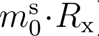) is reduced in *clinical* AD and is predictive of amnestic MCI to AD conversion (Duan et al., 2022; Giulietti et al., 2012; Makovac et al., 2018). Our results using an *unconstrained* qMT model, however, suggest that *R*_x_ is more sensitive to amyloid pathology than 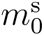 in *preclinical* AD. This finding raises an interesting possibility: if neurodegeneration causes reduced 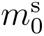 in advanced disease (discussed further in Section 4.2), *R*_x_ and 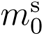 may each potentially be associated with the ‘A’ and ‘N’ axes of the A/T/N framework (Jack et al., 2018), respectively. This emphasizes the importance of separating these parameters in the unconstrained qMT model.

Contrary to our expectations of an increase with greater amyloid burden, we found no significant group differences in the macromolecular pool size 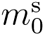, although a positive correlation was observed in the temporal lobe. One explanation could be a competing effect causing a simultaneous decrease in 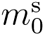, possibly due to concomitant neurodegeneration or a change in the interstitial load of water (for example, due to reduced fluid clearance (Tarasoff-Conway et al., 2015) or vascular leakage from reactive astrogliosis (Nakahara et al., 1999)). However, the latter hypothesis appears less likely given the lack of significant changes we would expect in the free water pool’s relaxation rates 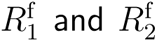 (Assländer, Mao, et al., 2024; Stanisz et al., 2004).

### 4.1 Limitations

Our preliminary study was designed to assess the utility of unconstrained qMT biomarkers for detecting Aβ accumulation and has several limitations. Firstly, a larger cohort is needed to verify our findings, control for the effect of nuisance variables (e.g., age, sex, race), and increase the statistical power to perform ROI or voxel-level analyses. Secondly, mechanistic studies are needed to elucidate the specific pathophysiological processes underpinning the observed changes in *R*_x_ and 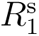, including potential covariates like blood-brain barrier breakdown (Tarasoff-Conway et al., 2015), temperature (Klegeris et al., 2006), or pH changes (Louie et al., 2008). For example, altered perfusion could explain the decreased 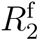 observed in subcortical structures and white matter. Temperature differences of even 1–2 *^◦^*C could explain the observed cortical differences in *R*_x_ and 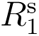 (Birkl et al., 2013). pH changes due to mitochondrial dysfunction (Wang et al., 2020) would decrease *R*_x_ and may have preceded cognitive decline in our cohort. Additionally, a post-mortem study of the relationship between 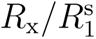 and neuritic plaque density would provide a stronger histopathological basis for our data. Lastly, our findings should be validated in preclinical (e.g., mouse) models of AD, as our unconstrained model differs from the existing literature primarily utilizing constrained qMT approaches (Bigot et al., 2014; Pérez-Torres et al., 2014; Praet et al., 2016).

### 4.2 Future Directions

Our proof-of-concept study was based on a prototype qMT sequence originally designed for the study of white matter. Future work will involve optimizing the sequence for the quantification of typical grey matter values using the same procedure described in Assländer, Mao, et al. (2024). Further, the qMT maps exhibited artifacts particularly affecting ROIs in the frontal and parietal lobes, which could explain their relative lack of significant effects compared to the temporal lobe. One likely source of artifacts in the frontal and parietal lobes, especially considering the employed radial k-space trajectory, is off-resonance effects arising from the chemical shift of subcutaneous fat with respect to water. Future work will involve correcting for fat-water chemical shift-related artifacts.

The qMT sequence has an effective 1.24 mm isotropic resolution. Future work will explore the potential advantages of this high resolution (as compared to PET) in the study of finer structures relevant to AD, including the hippocampus, cortex, and brainstem. For the latter, decreased neuromelanin and degeneration of the locus coeruleus in the pons has been associated with increased levels of CSF Aβ (Betts et al., 2019) and vulnerability to the occurrence of neurofibrillary tangles (Braak et al., 2011) in early-stage AD. As suggested by Trujillo et al. (2019), qMT could potentially be used to detect decreased neuromelanin in the locus coeruleus.

Intriguingly, our data in the subcortical grey matter shows a significant decrease in both of the free pool’s relaxation rates 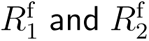 for Aβ+ subjects, specifically in the hippocampus and thalamus (Figure 4 and Supporting Table S3). One possible explanation for these differences is inflammation (Leng & Edison, 2021), which is known to cause decreases in the relaxation rates (Stanisz et al., 2004). Iron is known to accumulate and co-localize with Aβ in the hippocampal subiculum based on post-mortem AD tissue samples (Madsen et al., 2020; Smith et al., 1997; Zeineh et al., 2015), and concentrations of the ferrous form, which causes oxidative stress, have been proposed to increase with microglial-driven inflammation (Tran et al., 2022), which may link Aβ pathology with subsequent neurodegeneration (Leng & Edison, 2021). However, our previous data shows that both 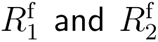 in the unconstrained qMT model are highly positively correlated with iron concentration (Assländer, Mao, et al., 2024). Future studies could incorporate quantitative susceptibility-weighted imaging (Ayton et al., 2017; Yedavalli et al., 2021) to verify possible hippocampal changes in iron content within the preclinical AD population.

Unconstrained qMT should also be extended to study injury in white matter areas (Makovac et al., 2018) where 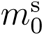 was previously demonstrated to be closely associated with myelin content (Janve et al., 2013; Thiessen et al., 2013), but amyloid PET has significant off-target binding (Chapleau et al., 2022). Additionally, we hypothesize that cortical grey matter neurodegeneration in clinical AD would also be reflected by reduced 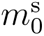. This motivates further investigation into which unconstrained qMT parameters might be sensitive to disease progression. Under the model of amyloid accumulation and progressive neurodegeneration as temporally displaced processes (Jack et al., 2010), qMT’s potential sensitivity to both ‘A’ and ‘N’ axes (Jack et al., 2018) could improve the specificity for and monitoring of AD stage. Further investigation is needed to establish unconstrained qMT’s sensitivity to patients with more advanced disease and specific associations with the neurodegenerative axis of the A/T/N framework. Along similar lines, more work is also needed to clarify whether qMT parameters are sensitive to aggregates of tau proteins (like neurofibrillary tangles).

qMT—which combines high-resolution anatomical imaging and amyloid sensitivity in a single exam—may also be useful for the longitudinal monitoring of therapeutic response in clinical trials of disease-modifying drugs. In particular, emerging anti-amyloid immunotherapeutics are increasingly being studied in preclinical patient populations (Yadollahikhales & Rojas, 2023); e.g., in the AHEAD 3-45 study (NCT04468659). While the qMT effect sizes are substantially smaller than those of amyloid PET, qMT is more amenable to screening, longitudinal imaging, and multi-center studies by virtue of being implementable on existing MRI scanners. One important consideration also relevant for clinical trials is the detection of amyloid-related imaging abnormalities (ARIA), which are used as a stopping criterion at the individual and study level according to FDA guidelines. We anticipate that both forms of ARIA are likely detectable in the free water relaxation rates 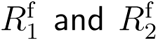 (Assländer, Mao, et al., 2024), though it remains to be seen if unconstrained qMT adds value to the assessment of ARIA in comparison to standard MRI protocols.

## 5 Conclusion

Our study is the first to utilize an unconstrained qMT model to compare qMT parameters directly to an accepted measure of amyloid burden (amyloid PET) in pre-symptomatic individuals on the Alzheimer’s disease spectrum. We show that the magnetization exchange rate and the semi-solid spin pool’s longitudinal relaxation rate are potential biomarkers of amyloid beta accumulation. qMT is a minimal addition to routinely used conventional MRI that may enable the detection of amyloid accumulation without requiring contrast agents or radiotracers. Future studies are needed to establish a potential role for qMT in monitoring disease progression and the response to therapy.

## Data and Code Availability

The qMT parameter maps, MPRAGE, and FBB PET SUV images for all participants are available at https://zenodo.org/records/11479570 (doi: 10.5281/zenodo.11479570). The source image reconstruction code used for the qMT data is available as a *Julia* package on Github at https://github.com/ JakobAsslaender/MRFingerprintingRecon.jl. For the presented data, we used package v0.7.0 with Julia v1.10.0.

## Author Contributions

AM: Conceptualization, data curation, formal analysis, funding acquisition, investigation, methodology, project administration, software, validation, visualization, writing – original draft, and writing – review & editing. SF: Methodology, software, and writing – review/editing. EM: Software and writing – review/editing. AVM: Funding acquisition, resources, and writing – review & editing. HR: Methodology, resources, supervision, and writing – review & editing. JA: Conceptualization, funding acquisition, methodology, software, supervision, and writing – review & editing.

## Data Availability

N/A

## Acknowledgements

The authors are grateful to David H. Salat, Ryn Flaherty, and Yu Veronica Sui for their helpful pointers regarding the use of the Freesurfer package, as well as Zena Rockowitz for her assistance with subject recruitment.

## Supplementary Material

Supplementary material is provided online.

