## Supplementary Material for "Sensitivity of unconstrained quantitative magnetization transfer MRI to Amyloid burden in preclinical Alzheimer’s disease"

### Supplementary Data for: Sensitivity of unconstrained quantitative magnetization transfer MRI to Amyloid burden in preclinical Alzheimer's disease

Andrew Mao,[a,b,c*] Sebastian Flassbeck,[a,b] Elisa Marchetto,[a,b], Arjun V. Masurkar,[d,e,f] Henry Rusinek,[a,b,d,g], Jakob Assländer[a,b]

[a]Bernard and Irene Schwartz Center for Biomedical Imaging, Department of Radiology,

New York University Grossman School of Medicine, New York, NY, USA

[b]Center for Advanced Imaging Innovation and Research (CAI$^2$R), Department of Radiology,

New York University Grossman School of Medicine, New York, NY, USA

[c]Vilcek Institute of Graduate Biomedical Sciences, New York University Grossman School of Medicine, New York, NY, USA

[d]Alzheimer's Disease Research Center, Center for Cognitive Neurology,

New York University Grossman School of Medicine, New York, NY, USA

[e]Department of Neurology, New York University Grossman School of Medicine, New York, NY, USA

[f]Department of Neuroscience and Physiology, New York University Grossman School of Medicine, New York, NY, USA

[g]Department of Psychiatry, New York University Grossman School of Medicine, New York, NY, USA

June 17, 2024

| Cortical Lobe | FBB SUVR | | $m_0^s$ | | $R_x$ | | $R_1^s$ | | Thickness | |
|---|---|---|---|---|---|---|---|---|---|---|
| | $p$ | $g$ | $p$ | $g$ | $p$ | $g$ | $p$ | $g$ | $p$ | $g$ |
| Frontal Ctx | $2{\cdot}10^{-8}$ | 3.49 | 0.200 | 0.49 | 0.104 | −0.62 | 0.039 | −0.78 | 0.851 | 0.10 |
| Parietal Ctx | $2{\cdot}10^{-8}$ | 3.69 | 0.305 | 0.41 | 0.087 | −0.60 | 0.019 | −0.81 | 0.215 | 0.45 |
| Temporal Ctx | $2{\cdot}10^{-8}$ | 2.32 | 0.124 | 0.63 | 0.022 | −0.86 | 0.012 | −0.94 | 1.00 | 0.01 |
| Occipital Ctx | $2{\cdot}10^{-8}$ | 2.46 | 0.305 | 0.44 | 0.884 | 0.01 | 0.249 | −0.48 | 0.232 | 0.28 |
| Cort. Sig. | $2{\cdot}10^{-8}$ | 2.83 | 0.267 | 0.34 | 0.095 | −0.76 | 0.087 | −0.63 | 0.983 | −0.03 |

Supporting Table S1: Summary of $p$ (for the non-parametric Mann-Whitney $U$ test) and Hedge's $g$ values for the measures and ROIs shown in Figure 3. "Cort. Sig." refers to the "signature of AD-related cortical thinning" described in Dickerson et al. (2011). Insignificant $p$-values ($p > 0.05$) and their associated $g$'s are shaded in grey.

| Cortical Lobe | $R_1^f$ | | $R_2^f$ | | $T_2^s$ | |
|---|---|---|---|---|---|---|
| | $p$ | $g$ | $p$ | $g$ | $p$ | $g$ |
| Frontal Ctx | 0.692 | 0.23 | 0.573 | −0.17 | 0.465 | −0.33 |
| Parietal Ctx | 0.692 | 0.11 | 0.950 | −0.18 | 0.723 | −0.23 |
| Temporal Ctx | 0.215 | 0.37 | 0.285 | 0.31 | 0.662 | 0.12 |
| Occipital Ctx | 0.950 | −0.05 | 0.465 | −0.32 | 0.787 | −0.27 |
| AD-signature | 0.787 | 0.11 | 0.851 | 0.04 | 0.185 | −0.49 |

Supporting Table S2: Summary of $p$ and $g$ values for the remaining qMT parameters not shown in Figure 3. Insignificant $p$-values ($p > 0.05$) and their associated $g$'s are shaded in grey.

| ROI | FBB SUVR | | $m_0^s$ | | $R_1^f$ | | $R_2^f$ | | Volume | |
|---|---|---|---|---|---|---|---|---|---|---|
| | $p$ | $g$ | $p$ | $g$ | $p$ | $g$ | $p$ | $g$ | $p$ | $g$ |
| Hippocampus | 0.001 | 1.37 | 0.146 | −0.28 | 0.005 | −0.83 | 0.036 | −0.87 | 0.107 | −0.66 |
| Amygdala | $1{\cdot}10^{-4}$ | 1.55 | 0.387 | 0.37 | 0.833 | 0.01 | 0.526 | −0.26 | 0.076 | −0.69 |
| Thalamus | 0.004 | 1.25 | 0.552 | −0.26 | 0.009 | −1.03 | 0.040 | −0.78 | 0.863 | −0.09 |
| Caudate | $4{\cdot}10^{-8}$ | 2.34 | 0.064 | −0.63 | 0.307 | −0.48 | 0.158 | −0.41 | 0.255 | −0.43 |
| Putamen | $9{\cdot}10^{-9}$ | 2.45 | 0.408 | −0.39 | 0.064 | −0.64 | 0.289 | −0.43 | 0.924 | −0.06 |
| Pallidum | 0.06 | 0.78 | 0.017 | −0.95 | 0.008 | −1.00 | 0.195 | −0.60 | 0.346 | −0.39 |
| Global WM | $4{\cdot}10^{-7}$ | 2.52 | 0.107 | −0.61 | $7{\cdot}10^{-5}$ | −1.54 | 0.010 | −1.21 | 0.107 | −0.72 |

Supporting Table S3: Summary of $p$ and $g$ values for the measures and ROIs shown in Figure 4. Insignificant $p$-values ($p > 0.05$) and their associated $g$'s are shaded in grey.

| ROI | $R_x$ | | $R_1^s$ | | $T_2^s$ | |
|---|---|---|---|---|---|---|
| | $p$ | $g$ | $p$ | $g$ | $p$ | $g$ |
| Hippocampus | 0.058 | 0.72 | 0.255 | 0.30 | 0.064 | 0.53 |
| Amygdala | 0.526 | 0.23 | 0.272 | 0.45 | 0.170 | 0.49 |
| Thalamus | 0.170 | −0.64 | 0.255 | 0.46 | 0.010 | 1.0 |
| Caudate | 0.326 | −0.40 | 1.00 | 0.07 | 0.985 | 0.11 |
| Putamen | 0.863 | −0.18 | 0.833 | −0.23 | 0.289 | 0.53 |
| Pallidum | 0.224 | −0.49 | 0.716 | 0.04 | 0.346 | 0.32 |
| Global WM | 0.774 | −0.04 | 0.687 | −0.09 | 0.026 | 0.76 |

Supporting Table S4: Summary of $p$ and $g$ values for the remaining qMT parameters not shown in Figure 4. Insignificant $p$-values ($p > 0.05$) and their associated $g$'s are shaded in grey.
